# The clinical need for clustered AChR cell-based assay testing of seronegative MG

**DOI:** 10.1101/2022.03.07.22272014

**Authors:** Gianvito Masi, Yingkai Li, Tabitha Karatz, Minh C. Pham, Seneca R. Oxendine, Richard J. Nowak, Jeffrey T. Guptill, Kevin C. O’Connor

**Author notes:** **Corresponding Author:** Kevin C. O’Connor PhD, Yale School of Medicine, 300 George Street Room 353J, New Haven, CT 06511. These authors contributed equally to this work (Richard J. Nowak, Jeffrey T. Guptill, Kevin C. O’Connor).

## Abstract

Trial eligibility in myasthenia gravis (MG) remains largely dependent on a positive autoantibody serostatus. This significantly hinders seronegative MG (SNMG) patients from receiving potentially beneficial new treatments. In a subset of SNMG patients, acetylcholine receptor (AChR) autoantibodies are detectable by a clustered AChR cell-based assay (CBA). Of 99 SNMG patients from two academic U.S. centers, 18 (18.2%) tested positive by this assay. Autoantibody positivity was further validated in 17/18 patients. In a complementary experiment, circulating AChR-specific B cells were identified in a CBA-positive SNMG patient. These findings corroborate the clinical need for clustered AChR CBA testing when evaluating SNMG patients.

## Introduction

In clinical practice, autoantibody (Ab) testing is the cornerstone of the diagnosis and management of myasthenia gravis (MG). Indeed, a positive result translates into two practical consequences, as it confirms the clinical suspicion of MG and directly informs therapeutic strategies. Radioimmunoassay (RIA) is the current gold standard for the detection of Abs binding to the acetylcholine receptor (AChR) or muscle-specific tyrosine kinase (MuSK), which are found respectively in ∼80% and 1-10% of all MG patients.^1^ Despite the identification of new autoantigens in MG, such as low-density lipoprotein receptor-related protein 4 and agrin, Abs are still undetectable in ∼10% of generalized MG cases.^1-3^ This group of patients, defined as “seronegative” (SN), raises two major unmet issues which continue to challenge existing clinical standards. First, given a lack of detectable Abs, confirmation of a diagnosis of SNMG is more difficult to establish. The availability of advanced neurophysiological testing, such as repetitive nerve stimulation and single-fiber electromyography, is limited in some regions. In addition, abnormal neuromuscular transmission on neurophysiological testing, when considered alone, can be insufficient, as it does not discriminate between autoimmune MG and congenital myasthenic syndromes, the latter of which are not appropriate for immunosuppressive therapy.^4,5^ Second, promising new drugs for MG are in the pipeline, and others are currently in clinical trials, but therapeutic eligibility remains largely dependent on a positive Ab serostatus. This poses a further problem for equitable access to healthcare, as the majority of such treatments are considered off-label for SNMG patients, and payers typically limit access only to seropositive patients.

In 2008 Leite et al. reported that a cell-based assay (CBA) in which AChRs are densely clustered on the cell membrane through rapsyn could detect AChR Abs in 66% of a RIA-based AChR and MuSK Ab-negative MG cohort.^6^ Later studies have confirmed this finding in variable proportions of SNMG patients.^7-9^ Notwithstanding the increased sensitivity of clustered AChR CBAs in detecting AChR Abs, the technical expertise and specialized equipment required by these assays have limited their use to research institutions. Still, a systematic employment could expedite the diagnosis of MG when evaluating SNMG patients, granting them potential eligibility for current and future treatments. Data on the frequency of SNMG patients with CBA-positive AChR Abs in the United States are lacking. To this end, we performed a clustered AChR CBA to screen a large cohort of SNMG patients evaluated by two academic centers in the U.S. Furthermore, we validated the CBA results with different approaches to confirm the presence of AChR Abs and AChR-specific B cells.

## Methods

This study was approved by Yale and Duke Universities’ Institutional Review Board. Patients or their legally authorized representative consented to participate in the study. Serum or plasma samples of SNMG patients were sourced from the Yale and Duke Myasthenia Gravis Clinics’ biorepositories (NCT03792659). SNMG was diagnosed based on clinical and electromyographic criteria, and the lack of AChR and MuSK Abs detected by RIA. Demographic and clinical data were obtained by chart review. All specimens were tested at 1:20 dilution through a live clustered AChR CBA using flow-cytometry, as previously described.^10^ A human recombinant monoclonal antibody specific for AChR, mAb 637,^11^ serum samples from RIA-based AChR Ab-positive MG patients and healthy subjects were included as test controls. Positive CBA results at screening were further validated by two parallel approaches. First, CBA-positive samples underwent IgG purification using Protein G Sepharose 4 Fast Flow beads (GE Healthcare Life Sciences) according to the manufacturer’s instructions before being re-tested at a concentration of 300 µg/ml by a subsequent CBA. In a second approach, an additional CBA was performed with serial sample dilutions. A novel B cell culturing method, reported elsewhere,^12,13^ was employed on an exploratory basis to detect AChR-specific B cells in the peripheral blood mononuclear cells of SNMG patients.

## Results

All specimens were negative for AChR and MuSK Abs by RIA. From a total of 99 SNMG patients, 18 (18.2%) tested positive by clustered AChR CBA (**Figure, A**). In this group, median age at MG onset was 44 years (range 13-79), and 4/18 (22.2%) were female. At the time of blood sample collection, Myasthenia Gravis Foundation of America class was ≥ II in 7/18 (38.8 %) patients. In 17/18 patients with a positive CBA result at screening, the presence of AChR Abs was confirmed by both validation methods (**Figure, B-C**). To obtain further evidence of a positive or negative AChR Ab status assessed through CBA, we sought to detect circulating AChR-specific B cells in SNMG patients. To that end, we tested culture media from stimulated, blood-derived B cells of 4 SNMG patients, one of whom tested positive by the clustered AChR CBA. AChR-specific B cells were identified in the SNMG patient with a positive CBA result, but not in the other three who were CBA-negative (**Figure, D**).

**Figure.**
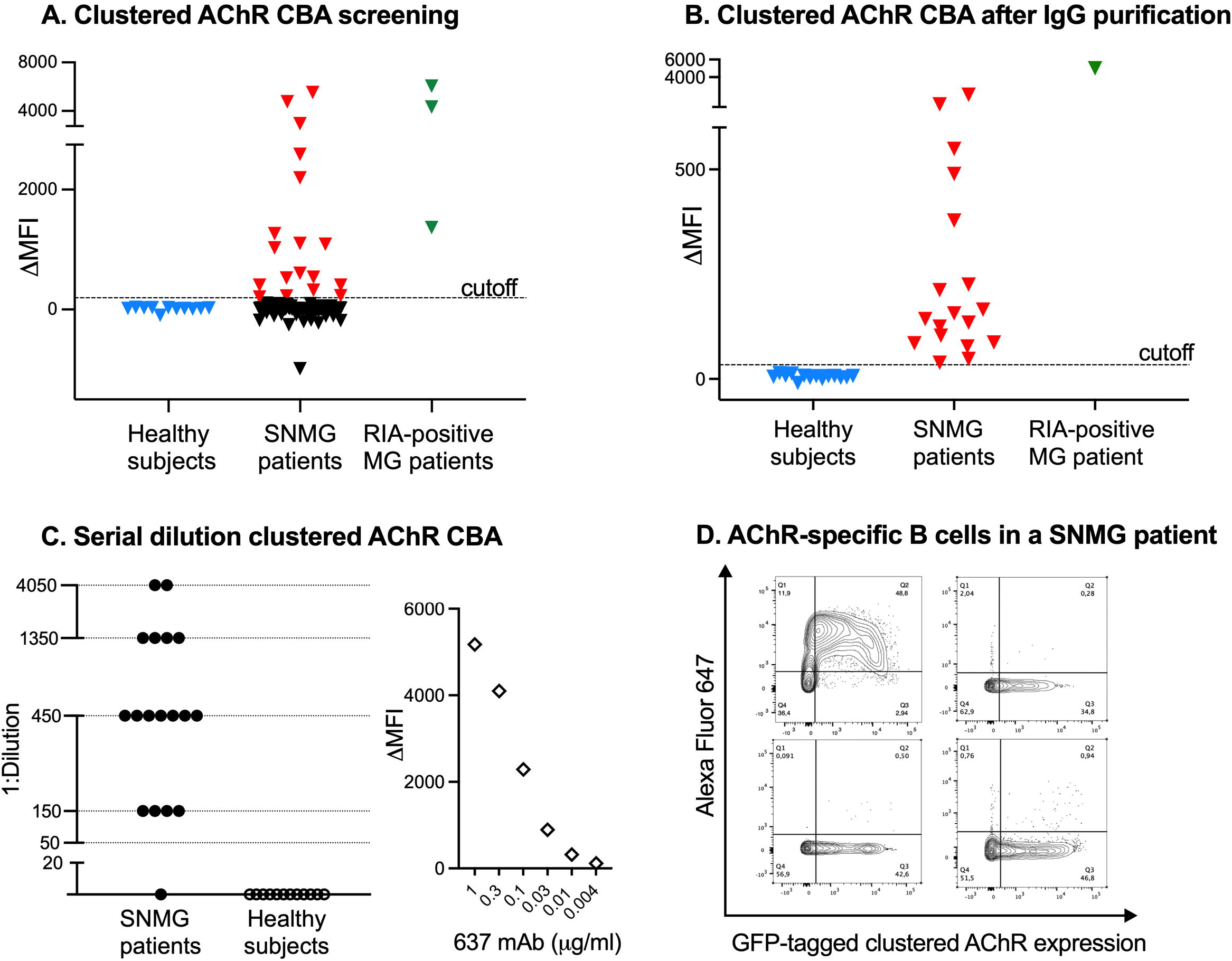
**(A) Scatter plot of clustered AChR CBA screening results by flow cytometry.** Serum or plasma samples from SNMG patients (n=99), healthy subjects (n=11) and RIA-based AChR Ab-positive MG patients (n=3) were tested at 1:20 (screening dilution) by a live flow cytometry-based clustered AChR CBA. HEK293T cells were transfected with plasmids encoding the adult AChR subunits (2α, β, δ, and ε) and green fluorescent protein (GFP)-tagged rapsyn. Alexa Fluor 647-conjugated anti-human Fcγ was used as a secondary antibody to detect IgG binding to the AChR transfected cells. For every sample tested, the median Alexa Fluor 647 fluorescence intensity (MFI) was measured in two cell populations: GFP-positive (high AChR expression) and GFP-negative HEK293T cells (low or no AChR expression). The difference between the two MFIs was calculated (ΔMFI). Each symbol (triangle) is the mean of duplicate experiments. The positivity cutoff was defined as the mean ΔMFI of healthy subjects plus 5 standard deviations. **(B) Clustered AChR CBA results after IgG purification or (C) with serial sample dilutions**. CBA-positive SNMG samples at screening (n=18/99, 18.2%), serum samples from healthy subjects (n=14), and one RIA-based AChR Ab-positive serum sample underwent IgG purification before being re-tested by a further CBA (each triangle is the mean of triplicates). The positivity cutoff was defined as the mean ΔMFI of healthy subjects plus 5 standard deviations. In a different validation approach (panel C, left), the same positive SNMG samples at screening (n=18) were tested by another CBA at 1:20 dilution plus 5 additional 3-fold dilutions (1:50-1:4050). Each black dot represents a SNMG sample at the maximum positive dilution. Data points with a value of y= 0 were negative at 1:20 dilution. As positive control (panel C, right), AChR-specific mAb 637 was tested on the same assay run at serial dilutions, starting from 1 µg/ml plus 5 additional 3-fold dilutions. **(D) Identification of circulating AChR-specific B cells in a SNMG patient**. After B cell differentiation into Ig-producing B cells, cell-culture supernatants were tested using the flow cytometry-based clustered AChR CBA. AChR IgG were detected (upper-left representative flow plot) in the culture supernatant from a SNMG patient with a positive CBA, indicating the presence of circulating AChR-specific B cells and further substantiating the positive AChR Ab status. In contrast, AChR-specific B cells were not detected in 3 other SNMG patients with a negative CBA (upper-right and bottom representative flow plots).

## Discussion

By performing a live flow cytometry-based clustered AChR CBA, AChR Abs were detected in approximately one-fifth of our U.S. SNMG cohort. Furthermore, we employed a method that allows for *in vitro* differentiation of B cells into antibody-secreting cells, through which we identified circulating AChR-specific B cells in a CBA-positive SNMG patient. This approach enabled us to substantiate a positive AChR Ab status first determined by CBA. Because of the exploratory nature of our study design, additional research is needed to investigate the generalizability of this finding to other CBA-positive SNMG patients.

These data nonetheless corroborate the clinical need to implement clustered AChR CBA testing in the evaluation of SNMG patients. The technical difficulties posed by setting up in-house live CBAs can be overcome by the employment of fixed CBAs, which are easier to perform and do not require cell culture facilities. Notably, diagnostic kits with fixed transfected cells are already employed in many centers and allow for widespread testing in suspected cases of other autoimmune neurological conditions such as autoimmune encephalitis and neuromyelitis optica.^14,15^ For some antigens such as myelin oligodendrocyte glycoprotein, comparative studies have shown a superior accuracy of live CBAs compared to fixed ones, in which fixation is likely to alter the protein conformation on the cell membrane, thus reducing test sensitivity.^16^ In the case of clustered AChR CBAs, prospective studies are needed to compare the diagnostic performance between live and fixed methodologies. In a recent study, a commercially available fixed AChR CBA had excellent specificity for MG comparable to RIA, as well as 4% higher sensitivity, suggesting that this assay could even replace RIA as the gold standard Ab test for MG.^17^ Another advantage of fixed (and live) CBAs is that they do not require radioactive agents used in RIA. Compared to the highly standardized RIA, however, the interpretation of CBA results is not always straightforward, especially when a visual scoring system by microscopy is used and the signal approaches the subjective cutoff. Flow cytometry CBAs, unlike microscopy CBAs, have the advantage of yielding quantitative results, but their technical complexities may restrict their use to reference laboratories and academic centers.

Although test optimization is still needed for CBAs, they could connect a greater proportion of MG patients with current and future Ab-tailored treatment strategies such as thymectomy, complement inhibitors or neonatal Fc receptor blockers. Furthermore, by identifying Abs that remain undetected by RIA, CBAs might help to recognize cases which are truly seronegative for known Abs. This approach could aid in selecting those patients for whom further investigations, including genetic testing, may rule out MG-mimicking conditions. At the same time, it would narrow the subset of MG patients with Abs that bind to potential but still unidentified muscle autoantigens, which represent a worthy focus of future research.

## Data Availability

Anonymized data will be shared on reasonable request from qualified investigators, and upon completion of appropriate materials transfer agreements between institutions.

## Acknowledgments

This work was supported by the Targeted Research Project for Seronegative Myasthenia Gravis award from the Myasthenia Gravis Foundation of America (MGFA). Additional support to Dr. Kevin O’Connor was provided by the National Institute of Allergy and Infectious Diseases of the NIH under award numbers R01-AI114780 and R21-AI164590.

## Author Contributions

KCO, JTG and RJN: conception and design of the study; GM, YL, TK, MCP, SRO, RJN, JTG, and KCO: acquisition and analysis of data; GM: initial development and drafting of the manuscript and figure; GM, YL, TK, MCP, SRO, RJN, JTG, and KCO: editing of the manuscript.

